# Optimizing ventilation cycles to control airborne transmission risk of SARS-CoV2 in school classrooms

**DOI:** 10.1101/2020.12.19.20248493

**Authors:** Alessandro Zivelonghi, Massimo Lai

## Abstract

Open schools in winter in highly epidemic areas pose a controversial issue: ventilation of classrooms (an essential mitigation factor for airborne transmission) is expected to sensibly decrease due to outdoor temperatures getting colder and regulators going to allow less restrictive policies on windows closure. Fundamental questions to be addressed are therefore: to which extent can we contain airborne transmission risk in schools? what would be the optimal ventilation strategy during the cold season considering the fact that most schools are not provided with mechanical ventilation systems? To try answering these questions a risk model for airborne transmission of covid-19 in classrooms has been develped based on previous models for tubercolosis and influenza. The separate cases of infective student and infective teacher, as well as infective teacher with microphone are investigated. We explored 3500 different air ventilation cycles for different lesson+break times and carried out a numerical optimization of the risk function. Safety risk-zones for breaks and lessons durations were estimated combining the effect of surgical masks and optimal windows opening cycles.

## 1. Introduction

According to past and recent literature (e.g., Leung [2020], Morawska [2020-2], and Escombe [2007]), long distance oral transmission of infective diseases like Tuberculosis and Influenza in confined environments could be significantly reduced by proper ventilation. Although most of the relevant past literature has focused on the spread of tuberculosis or influenza in hospitals, the general principle of airborne transmission is valid and pertinent also for SARS-CoV2 in the context of school classrooms since these are enclosed settings which may host asymptomatic sources. As for mitigation strategies, indirect oral transmission of SARS-CoV2 is believed to be effectively curbed through the frequent opening of windows or by mechanical HVAC systems [Morawska 2020-2]. Very recent (although still unpublished) measurements of SARS-CoV2 air concentrations in ventilated and non-ventilated hospital rooms performed by the Italian Regional Environmental Agency “ARPA Piemonte”, confirm this statement [Repubblica 2021]. Compared to natural ventilation, mechanical ventilation systems, when adequately configured, can be equally or even more effective [Morawska 2020-2, ASHRAE 2020]. However, unlike hospitals, the vast majority of schools worldwide are not equipped with such systems and will not be, at least for the foreseeable future (including the 2020/2021 school year). In the real context of school classrooms, groups of students, which can potentially include tens of individuals, share the same premises for hours with potentially insufficient ventilation. This greatly increases the likelihood of coming into contact with virus-loaded aerosol droplets generated by even only one infective student or teacher in the same classroom. This issue is of concern also when social distancing is correctly implemented because of the volumetric nature of aerosol clouds. On the other hand, as suggested also by intuition, frequent natural ventilation could help dilute the viral load by allowing fresh air from the outside to enter classrooms. The extent to which natural ventilation combined with face masks, may reduce the airborne contagion risk in case of an infective student source, has been the subject of a preliminary numerical investigation published by one of the authors in October 2020 [Zivelonghi 2020]. In the present work we are extending such predictions presenting the infective teacher case while improving the general GN-model with the effective volume of dilution. Furthermore, we show a comprehensive mitigation strategy which include the novelties of numerical optimization of ventilation cycles and predictions for the reduction of group contagion risk through a voice amplification system at reasonable cost. The duration of lesson intervals (breaks) is one of the most critical parameters during winter, since only during breaks one can imagine to fully open the windows and dilute the viral load. Otherwise, complete windows opening during lesson time will drop the ambient temperature considerably and increase the risks of other diseases. This motivates a theoretical/computational framework aiming to optimize air change cycles in classrooms. In the present work, we explored 3500 different air ventilation cycles for different lesson+break times and carried out a numerical optimization of the risk function. Safety risk-zones for breaks and lessons durations were estimated combining the effect of surgical masks and optimal windows opening cycles. The strategy illustrated in the present analysis is based on natural ventilation, which is simple, efficient, and virtually free to implement. The same modelling framework, however, can be employed for mechanical ventilation cycles based on HVAC systems (since what counts is ultimately the air exchange rate, not its source).

## 2. Materials and Methods

### 2.1 GN-model with perfect mixing and face masks

The infection risk model used in the present analysis is essentially based on the Gammaitoni-Nucci (GN) model, which is adequate for confined, ventilated environments [Gammaitoni 1997]. This model is based on the assumption that newly-produced viral particles are instantly diluted over the whole environment volume (perfect-mixing) and that the emission rate parameter ER_q_ (emission rate, i.e. number of viral particles generated per hour by each infective subject) is known, at least as an averaged value over the exposure time 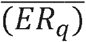 A correct estimation for the emission rate parameter ER_q_ is not trivial and mostly based on a semi-empirical approach combining viral load measurements from clinical trials and reverse engineering from observed outbreaks, assuming for ER_q_ averaged values selected from the variability ranges which arise from the following formula [Morawska 2009]. The “infective viral reduction rate” (IVRR) is the second parameter which enters the GN model and takes into account air ventilation (AER) and gravitational re-deposition (λ): *IVRR*=*AER*+λ. A λ value equal 0.5 vol/h was considered during the whole exposure time, on the basis of the half-life in air of the SARS-CoV-2 virus of about 1h as reported in [van Doremalen 2020]. According to the GN model, the risk of infection in a volume V, where one infective subject is present and the initial number of viral particles is n_0_ (which can be different from zero) is then given by the general formula:

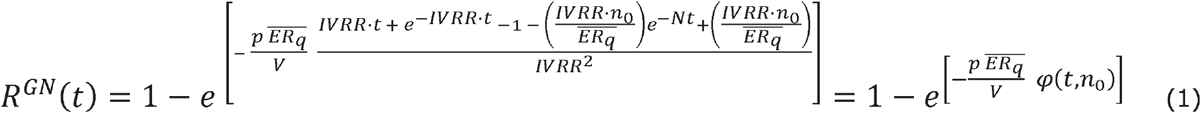

Where 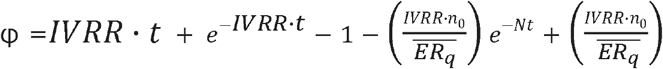

To account for the effect of PPE (personal protective equipment, in this case, face masks) in reducing both the number of viral particles generated by infective subjects, and also reducing the likelihood of inhalation of viral particles by exposed subjects, Eq (1)is extended introducing two scaling factors:

(1- f_out_), which represents the fractional reduction of the generated viral load, and

(1- f_in_), which represents the fractional reduction of inhaled viral load, under the assumption that all subjects are wearing a mask. Eq. (1) can then be rewritten as:

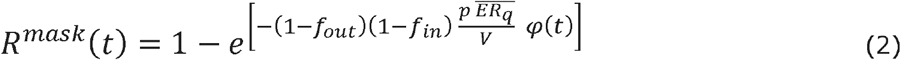

If masks are not being worn, *f*_*in*_ and *f*_*out*_ are both zero.

### 2.2 Influence of windows opening function on the infective virus removal rate (IVRR)

Windows opening implies a periodic activation of air exchanges per hour, which are supposed to occur only during lesson breaks. Therefore, the IVRR function in (3) become a periodic rectangular wave function over the full lesson time, with peaks influenced by higher values of the air exchange rate (AER) as due to complete windows opening (2vol/h) or mechanical ventilation (up to 10 vol/h). In this study we explore the influence on the classroom risk function of different cycles (and levels) of the AER within the IRVV function by changing the time duration of lecture and breaks. Fig. 2 report only two exemplary cycles (long and short windows opening cycles) taken as input for the results shown in Fig. 3-4, whereas optimization of the risk function shown in Fig. 5-6 considered 3500 different combination of t_brk_ and t_lec_ with a time resolution of 1 min which generated the same amount of different ventilation cycles. Another factor to be considered is the effective volume to be considered to dilute the aerosol viral cloud under the perfect mixing approximation. According to recent CFD simulations of aerosol cloud in classrooms [Abuhegazy 2020], aerosol particles from a student source would not be diluted over the entire volume even after a transient of 300s and the viral cloud volume during the first part of the emission transient would be negligible compared to V. For these reasons an effective lower volume V_*eff*_ = 0.85 V in equation (3) and (4) was considered for the present analysis.

In the present work, the GN-model is based on the concept of cumulative risk. In a scenario where the infective source is removed from the environment (e.g. student or teacher leaving the classroom), the ER_q_ parameter in Eq (1) and (2) would become zero. However, we can observe that even when ER_q_ =0 in (2), the infection risk 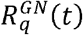 would remain greater than zero, because of the multiplicative factor cp *φq*(*t*) (and intuitively, due to the viral load already present in the environment).

Another underlying assumption, which is valid at least in Italian secondary schools and in many schools in EU countries, is that the susceptible subjects at the beginning (*S*_0_) remain always the same and do not vary over the course of the school day. For at least one infection to occur, the cumulative risk *R*_*c*_ (*t*)= *C*(*t*)/*S*_*0*_ must be greater than 1/ *S*_*0*_. Therefore, the condition for zero infections to occur over the total exposure time is:

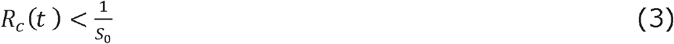

and not R_lec,i_ (t) < 1/S_0_.

Thus, at the end of a typical school day, one obtains the cumulative risk of infections with one infective source as given by:

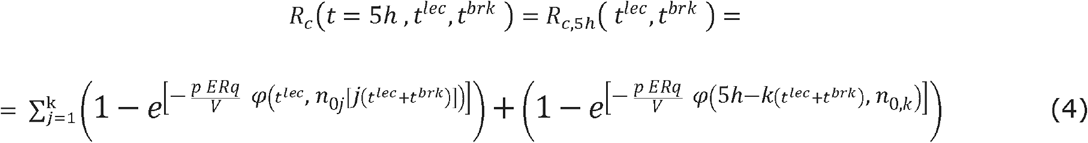

In equation (4), 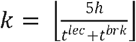 is the integer number of periods (lesson + break) before the last period. One may observe that the last lecture window could be less than *t*^*lec*^ in order not to overcome 5h of total lectures+breaks time. Equation (4) clearly shows that for a given classroom at the end of a school day, the only variables influencing the cumulative risk of airborne contagion (and the total number of infections) are the time duration of lectures (*t*^*lec*^) and the time duration of breaks (*t*^*brk*^), since the other model parameters (V, N, IVRR, p, ER_q_, f_out_, f_in_) can be considered either fixed or not controllable for a given classroom and a given local regulation. Two situations were separately considered. First, one positive student source remaining for 5 h in the same classroom but exiting it at each break interval (ER_q_ =0 at each break). Second, a positive teacher source supposed to remain for 2 hours only with higher ER_q_ levels. In fact, a high school teacher stays in the same classroom not more than two hours a day on average, moving to a new classroom afterwards. A python routine have been implemented to solve eq. (1-2) and (4) for both cases.

## 3. Results and Discussion

### 3.1. Optimum ventilation profiles and safety zones

Cumulative risk curves R_c_ (t) are shown in Fig. 1 and 2 as a function of two variable parameters as shown in Equation (4): *t*^*lec*^ (lecture time, when ventilation is turned off) and *t*^*brk*^ (duration of breaks, when ventilation is active). All surfaces were calculated for a typical classroom of volume 8 * 7 * 3 ≅ 170 m^3^. For the teacher case, higher levels of ER_q_ compared to the student case were required to account for a higher (average) speak activity. In all simulations the ER_q_ values employed were selected from published data [Buonanno 2020] for the case of standing person with normal (ER_q_ about 25 quanta h^-1^) and lowered vocal activity (average ER_q_ down to 15 quanta h^-1^). The latter case refers to a person speaking at moderate volume. This could be the case of a teacher speaking with a microphone connected to a voice amplification system. Even if staying in a classroom less than students (in average for only two hours), an infective teacher speaking most of that time will be a much greater viral source for airborne transmission and the corresponding risk curves will increase more steeply in the first 2h of exposure. After a teacher left the room, the ER_q_ in that room drops to zero, but the viral charge previously emitted by him/her will still be present for the next hours (although it will lower down after several ventilation cycles).

**Figure 1.**
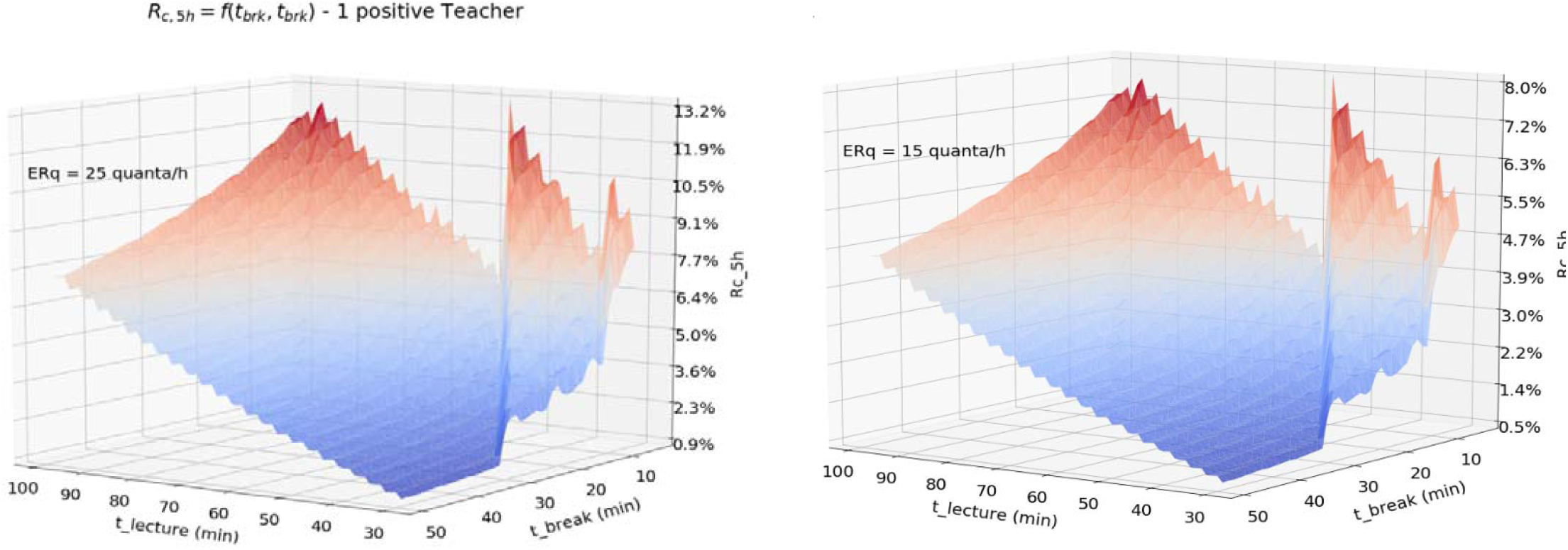
a,b) Cumulative risk function after 5h exposure as a function of lecture and break duration in the case of an infective teacher in classroom, for high (a) and low (b) emission rates. Hypotheses: classroom volume of 170 m3 and worse case for the effective wearing time of surgical masks (fin=0.15, fout=0.45).

**Figure 2.**
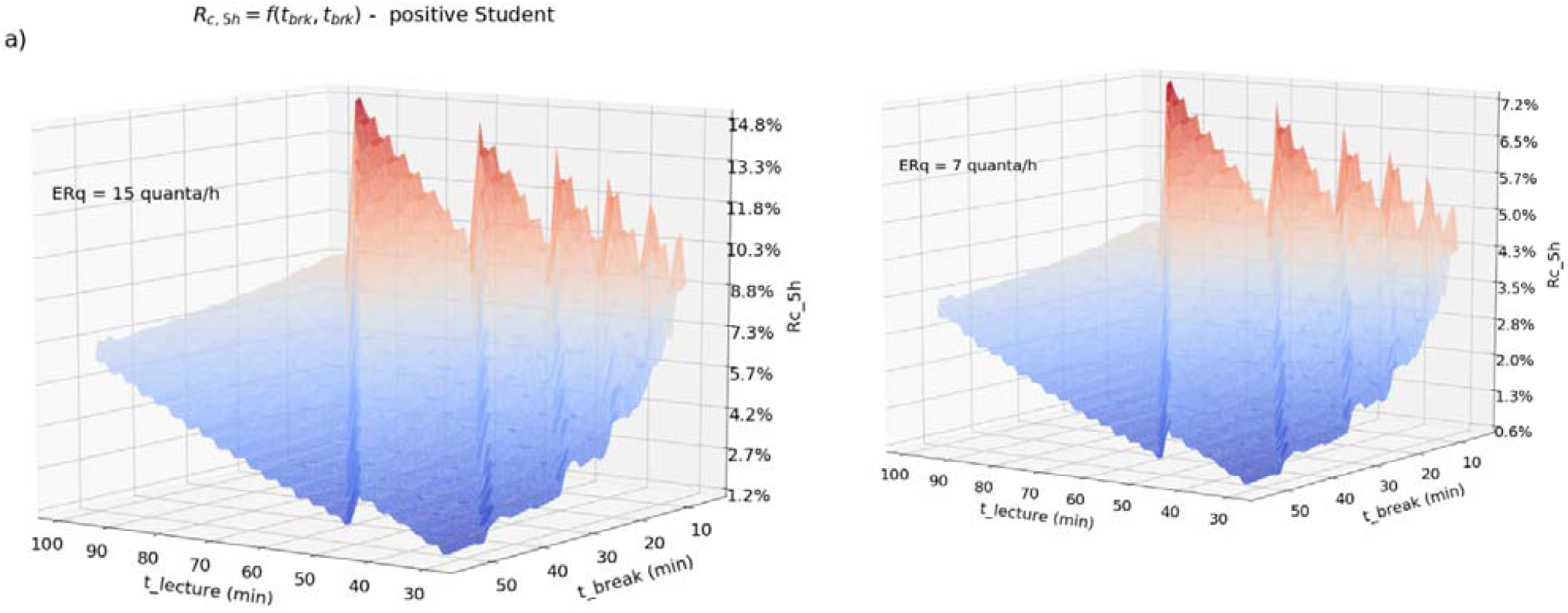
a,b) as in Fig. 1 but for the case of an infective student source.

**Table 1.** Model parameters and related value ranges. In brackets the controllable parameters.

| Parameters | Description | Units | Range |
| --- | --- | --- | --- |
| $t$ | duration exposure to infection | $h$ | 0-5 |
| $t_{brk}$ | breaks duration | min | [5-30] |
| $t_{lec}$ | lecture duration | min | [30-100] |
| $S$ | Number of susceptible persons | persons | |
| $C$ | Number of infected persons (contagions) | persons | |
| $R=C/S$ | infection risk | - | 0-100% |
| $ER_q$ | quanta emission rate by infective source | quanta $h^{-1}$ | [5-25] |
| $p$ | average inhalation flow | $m^3 h^{-1}$ | 0.6 |
| $V$ | classroom volume | $m^3$ | 170 |
| $V_{eff}$ | effective classroom volume | $m^3$ | 150 |
| $Q(t)$ | clean air inflow | $m^3 h^{-1}$ | 85-1700 |
| $AER(t)_{NV,MV}$ | air exchange rate provided by natural (NV) or mechanical ventilation (MV) | $h^{-1}$ | 0-2 (NV), 0-9.5 (MV) |
| $AER_{max}$ | max air changes per hour | $h^{-1}$ | 2 (NV), 9.5 (MV) |
| $\lambda$ | inactivation rate | $h^{-1}$ | 0.5 |
| $k$ | deposition rate | $h^{-1}$ | 0-0.25 |
| $IVRR(t)$ | infective virus removal rate $N+\lambda+k$ | $h^{-1}$ | 0-2.5 (NV), 0-10 (MV) |
| $n(t)$ | viral quanta in ambient at time $t$ | quanta | 0-50 |
| $n_0$ | viral quanta at time zero | quanta | 0 (>0 at each lesson reprise) |
| $f_{in}$ | mask reduction of inward viral load (susceptible person) | - | [15-30%] surgical |
| $f_{out}$ | mask reduction of outward viral load (source) | - | [45-90%] surgical |

As previously stated, a significant risk reduction could be obtained with more frequent ventilation. Thus, there is motivation for finding an optimal set of *t*^*lec*^,*t*^*brk*^ that would minimize *R*_*c,5h*_ (*t*^*lec*^,*t*^*brk*^), noting that only limited actions can be undertaken to vary the other exogenous parameters. Classroom volumes are usually fixed (except for the very few cases in which new spaces have been allocated in the current year). Second, acting on mask filter capabilities is also limited. That is, one may think it would improve the mask parameters f_out_ and f_in_ by requesting students to wear ffp2 masks all of the time, but this prescription for students younger than 18 years has been excluded by most regulators. In fact, wearing high-filtering masks in school contexts for long periods may expose students to other respiratory risks. The average pulmonary inflation p cannot be controlled in a lecture context and is reasonably known for human body. Finally, the critical emission rate parameter ER_q_ depends on actual habits/behaviors and is affected by uncertainties which were taken into account in the present study. In any case, both ER_q_ and p cannot be controlled by legislation (only ER_q_ for the teacher case could be lowered down by technical countermeasures, like the adoption of a microphone with amplifier). Instead, acting on ventilation (AER) and then on the directly linked IVVR is feasible in all schools and can be effective, as shown in this and other studies [Buonanno 2021]. On the other hand, maximizing AER in winter would require mechanical ventilation and operating HVAC systems, lacking in most schools. We can act only on windows opening for significant periods of time to increase the air exchange and dilution of the viral load. Therefore, an effective risk reduction pathway is the identification of a numerical optimum for the ventilation schedule as a function of time - AER_*opt*_*(t)* - which minimizes the cumulative risk within certain boundaries. The risk function to be minimized is z = *R*_*c,5h*_ (*t*^*lec*^,*t*^*brk*^) and is represented in Fig. 1 and Fig. 2 and the related contour plots of Fig. 3. As evident, a minimum exists at around (*t*^*lec*^,*t*^*brk*^) ≈ (30 min, 50*min*) since the z function is showing a clear monotonic behavior in the surrounding of the x=y direction. This would imply an unusual recommendation for most schools: class breaks should last longer than the classes themselves. This fact is eventually unsurprising, since opening windows during breaks implies a higher AER-level than during classes (Fig. A1 in Appendix). In addition, this fact is dictated by practical requirements. That is, with low outdoor temperatures (particularly in January–March in schools located in the northern hemisphere) and a concurrent risk for other seasonal diseases, it is only during “lesson breaks” with no students inside the classroom, that one could think to increase the AER substantially by fully opening the windows in a classroom.

**Figure 3.**
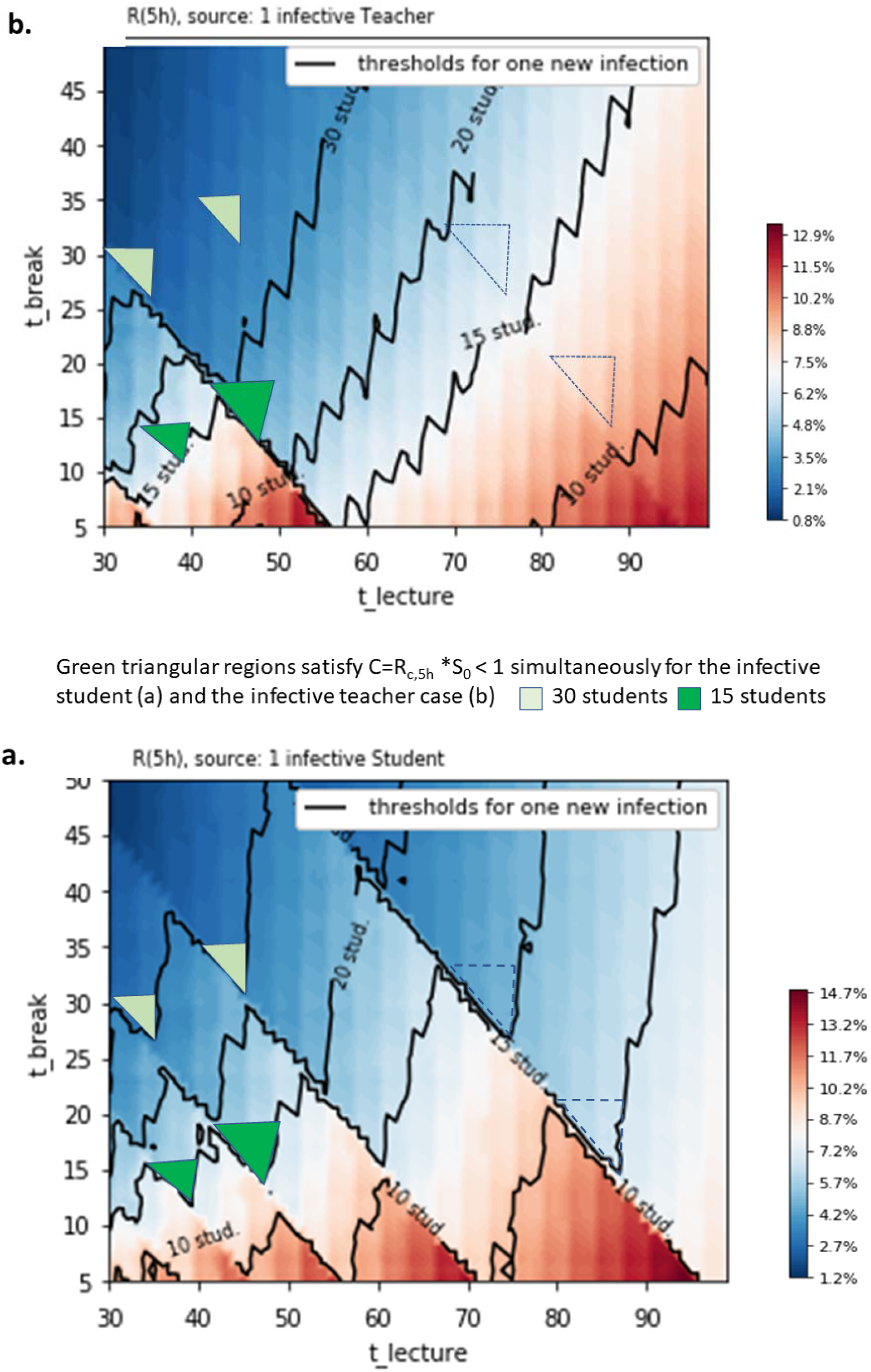
Contour plots of the cumulative risk function, R_c,5h_ (t^lec^,t^brk^) a) Positive student and b) positive teacher case. Continuous lines indicate “thresholds for a new infection” (C = 1) for different classroom groups. The regions they define are called “safety zones” for that group size. Green regions suggest values for t^lec^,t^brk^ which would be safe for the infective teacher as well for the infective student case, i.e. regardless of source type. Hypotheses are the same of Fig. 5.

However, since overly long breaks may also cause practical and organizational difficulties for schools, wider “safety zones” for the variation of *t*^*lec*^, *t*^*brk*^ are recommended in Fig. 6. These zones are defined below so-called “one-infection thresholds” (varying for different values of the number S0 of students per classroom). A certain combination of *t*^*lec*^,*t*^*brk*^ from the safety zone would thus provide sufficient ventilation and dilution of the viral load to lower the risk for new indirect infections below 1/S0. That is, no infections would be expected in a group of S0 susceptible individuals if the room were ventilated for an equal period of time 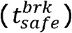 at every class break. Green regions suggest values for *t*^*lec*^, *t*^*brk*^ which would be safe for the infective teacher as well for the infective student case, i.e. regardless of source type. In order to achieve complete safety in a classroom of 30 students exposed for 5 hours, classes should last approximately 45 min but breaks should be very long (about 35 minutes). This would eventually turn into a complex or even unfeasible scheduling challenge for schools (only 3.75 lesson+break cycles would be possible within 5 hours). On the other hand, in a classroom of 15 students only, lessons could last 45-47 min and breaks easily reduced to 15 minutes, allowing 5 entire cycles.

## 4. Conclusions

Cumulative airborne risk is the key to understanding airborne infections of SARS-CoV-2 within classrooms. This originates from the cumulative nature of air saturation and viral aerosol formation. Furthermore, students and teachers exposed in schools for long periods of time to possible infection sources and standard sanitation/ventilation cycles during breaks cannot lower the residual viral load in their environment to zero. On the contrary, it has been shown that a small amount of this load would still be present in a classroom even after ventilation cycles at high N, with students leaving classrooms during breaks. On the other hand, an infective student re-entering a classroom would continue to emit viral quanta, and an infective teacher remaining in the room for a number of hours could emit sufficient viral quanta to indirectly infect other people, even after she/he exited the classroom. Although the dynamic single-zone model employed here contains some approximations, clear indications arises from this analysis. Firstly, windows in schools should be kept open most of the time to decrease the airborne risk to acceptable levels. Secondly, equipping teachers with microphone and voice amplification systems could be an additional effective mitigation strategy (in case of teachers as infective source).

Numerical results have indicated that windows should be fully (not partially) open during breaks and that breaks should possibly last as long as possible. However, since these recommendations may cause considerable organizational difficulties in schools and thermal discomfort in the winter season, compromise solutions are necessary. To prevent thermal discomfort and other seasonal diseases, schools could limit full natural ventilation during breaks. Therefore, specific safety regions for break and lecture durations (i.e., the parameters influencing ventilation profiles) for the different cases of infective teacher and infective student —have been identified. In this optimization, dilution and lowering of contagion risk were based entirely on natural ventilation cycles (i.e., opening windows) rather than on mechanical ventilation. Mechanical ventilation systems are expected at larger scale in future building upgrades but are not currently present in the vast majority of schools worldwide. Results of the minimization of the cumulative risk function indicate f.i. that alternating lectures of 45 min with breaks of 15 min would keep the contagion risk below the one-contagion-threshold in groups of 15 students, but for larger groups of 30 scholars much longer breaks of about 35 minutes would be necessary. Thus, as a general rule it may be wise, as suggested also by intuition, to have smaller class sizes to control airborne transmission in schools. Finally, it is once again remarked that only a combination of optimal air exchange cycles with protection masks worn by all individuals could lower the risk of airborne contagion in school classrooms to a desirable safety level of zero contagions in all conditions.

## Data Availability

all data are available running numerical python code which solve the equations presented in paper.

## Acknowledgment

The authors express gratitude to Prof. Buonanno and Prof. Morawska for insightful discussions.

## APPENDIX A

**Figure A1.**
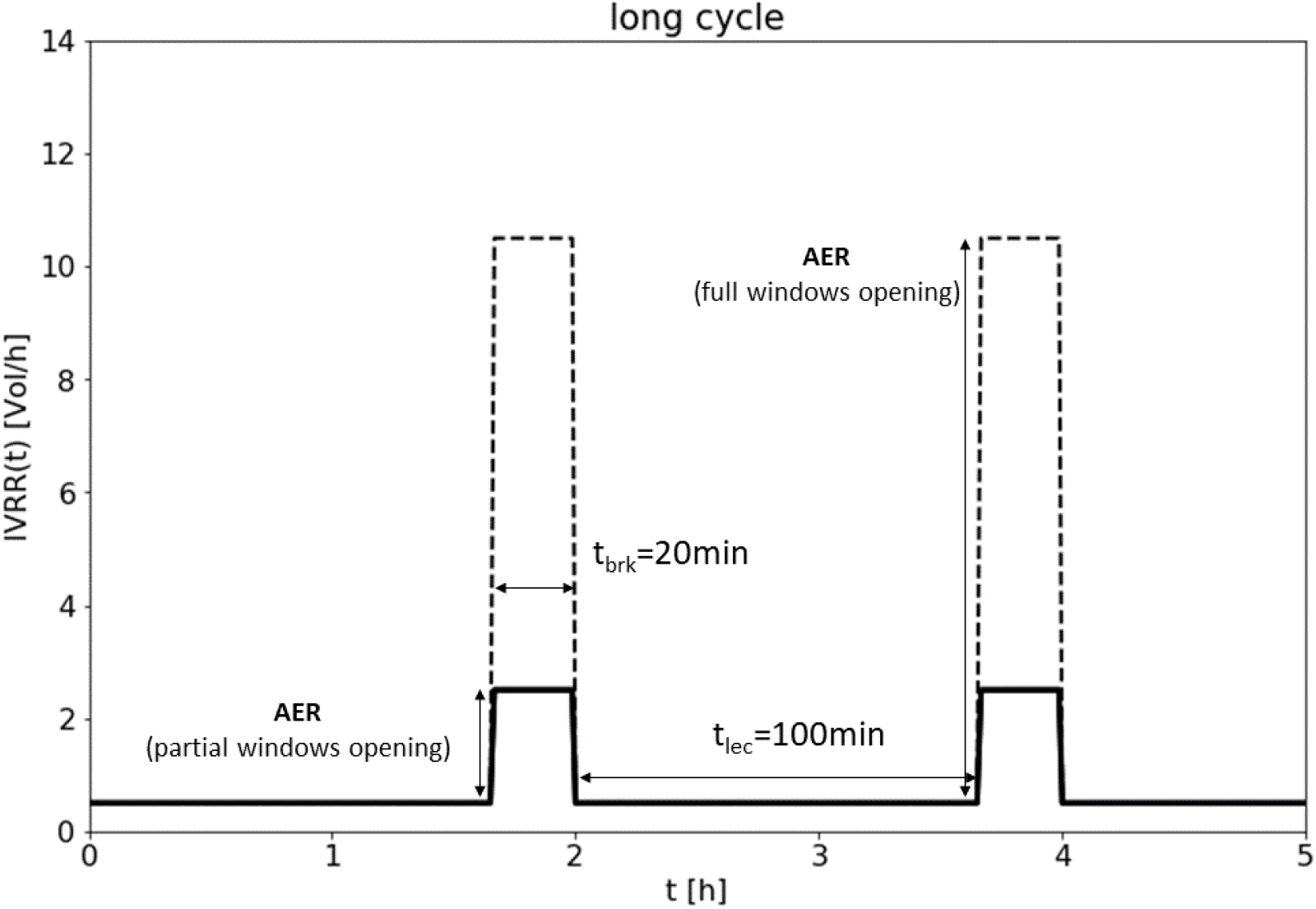
Example of two long ventilation cycles with different levels of maximum AER for windows opening

## SUPPLEMENTARY FIGURES

**Figure S1.**
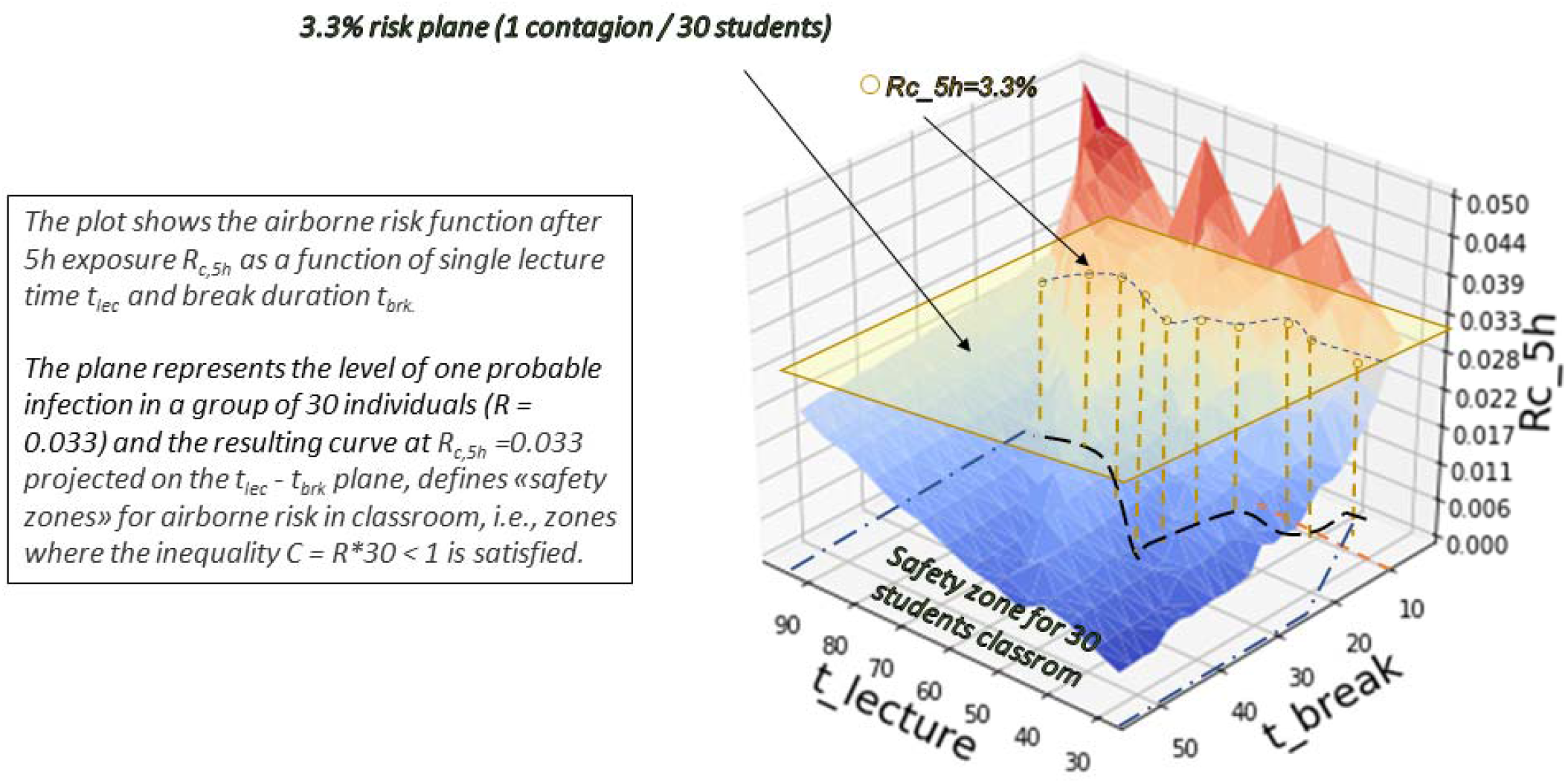
Definition of Safety Zone from the R_c,5h_ function

**Figure S2.**
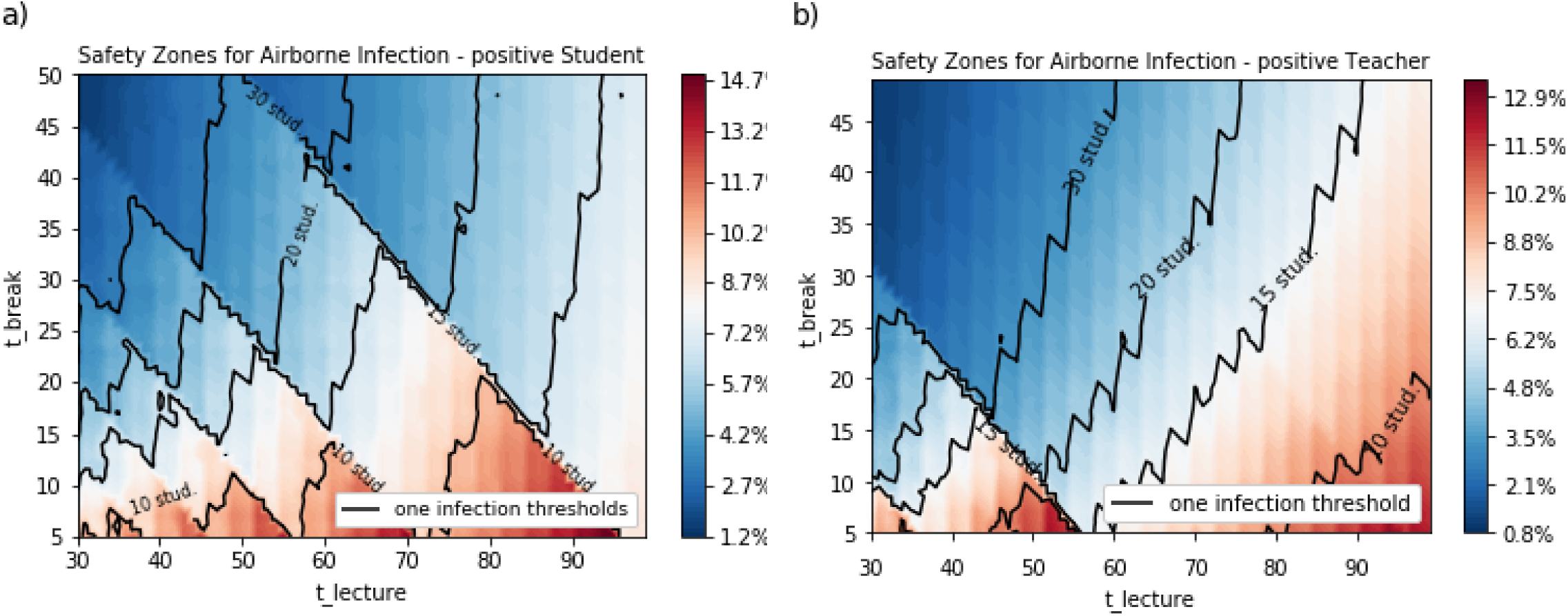
Figures 3a and 3b (without green triangles indicating optimality regions satistying both cases (for clarity)

## Bibliography

[Leung 2020] Leung, N.H.L., Chu, D.K.W., Shiu, E.Y.C. et al. Respiratory virus shedding in exhaled breath and efficacy of face masks. Nat Med 26, 676–680 (2020). https://doi.org/10.1038/s41591-020-0843-2

[Morawska 2020-1] Lidia Morawska, Donald K Milton, It is Time to Address Airborne Transmission of COVID- 19, Clinical Infectious Diseases, ciaa939, https://doi.org/10.1093/cid/ciaa939

[Qian 2020] Qian, H, Miao, T, Liu, L, Zheng, X, Luo, D, Li, Y. Indoor transmission of SARS-CoV-2. Indoor Air. 2020; 00: 1– 7. https://doi.org/10.1111/ina.12766

[Morawska 2020-2] Lidia Morawska, Julian W. Tang, William Bahnfleth, et al. How can airborne transmission of COVID-19 indoors be minimised? Environment International, Volume 142, 2020, 105832, ISSN 0160-4120, https://doi.org/10.1016/j.envint.2020.105832.

[Repubblica 2021] C. Zunino reporting on ARPA Piemonte study in cooperation with antiviral research laboratory “San Luigi Gonzaga”, Orbassano– “Virus in quali luoghi ce n’è di più. Inesistente all’ aperto, concentrazioni alte nelle case”. https://www.repubblica.it/scienze/2021/01/12/news/ora_si_potra_avvistare_il_covid_nell_aria-282183617/

[Allen 2020] Allen JG, Marr LC. Recognizing and controlling airborne transmission of SARS-CoV-2 in indoor environments. Indoor Air. 2020;30(4):557–558. doi:10.1111/ina.12697

[Wei 2020] Li Wei, Ji Lin, Xiaofei Duan, Wenzhi Huang, Xiaojun Lu, Juan Zhou, Zhiyong Zong Asymptomatic COVID-19 Patients Can Contaminate Their Surroundings: an Environment Sampling Study mSphere Jun 2020, 5 (3) e00442– 20; DOI: 10.1128/mSphere.00442-20

[Escombe 2007] Escombe AR, Oeser CC, Gilman RH, et al. Natural ventilation for the prevention of airborne contagion. PLoS Med. 2007; 4(2):e68. https://doi:10.1371/journal.pmed.0040068

[EN 16798] Energy Performance of Buildings - Ventilation for Buildings Part 7: Calculation Methods for the Determination of Air Flow Rates in Buildings Including Infiltration (Modules M5-5)

[Buonanno 2020 A] G. Buonanno, L. Stabile, L. Morawska. Estimation of airborne viral emission: Quanta emission rate of SARS-CoV-2 for infection risk assessment, Environment International, Volume 141,2020, 105794, ISSN 0160-4120, https://doi.org/10.1016/j.envint.2020.105794.

[Buonanno 2020 B] G. Buonanno, L. Stabile, L. Morawska. Quantitative assessment of the risk of airborne transmission of SARS-CoV-2 infection: Prospective and retrospective applications, Environment International, Volume 145, December 2020, 106112 ISSN 0160-4120, https://doi.org/10.1016/j.envint.2020.106112.

[AICARR 2020] AICARR - Protocollo per la riduzione del rischio da diffusione del SARS-CoV-2 nella gestione e manutenzione degli impianti (to be changed with corresponding ASHRAE protocol)

[Gammaitoni 1997] Gammaitoni L., Nucci M.C. 1997. Using a mathematical model to evaluate the efficacy of TB control measures. Emerging Infectious Disease, 3, 335–342.

[Knibbs 2011] Knibbs LD, Morawska L, Bell SC, Grzybowski P. Room ventilation and the risk of airborne infection transmission in 3 health care settings within a large teaching hospital. Am J Infect Control. 2011 Dec;39(10):866–72.. Epub 2011 Jun 12. PMID: 21658810 doi: https://doi.org/10.1016/j.ajic.2011.02.014

[Smereka 2020] Smereka, J., Ruetzler, K., Szarpak, L., Filipiak, K. J., & Jaguszewski, M. (2020). Role of mask/respirator protection against SARS-CoV-2. Anesthesia and analgesia, 10.1213/ANE.0000000000004873. Advance online publication. https://doi.org/10.1213/ANE.0000000000004873

[van Doremalen 2020] van Doremalen, N., Bushmaker, T., Morris, D.H., Holbrook, M.G., Gamble, A., Williamson, B.N., Tamin, A., Harcourt, J.L., Thornburg, N.J., Gerber, S.I., LloydSmith, J.O., de Wit, E., Munster, V.J., 2020. Aerosol and Surface Stability of SARS-CoV-2 as Compared with SARS-CoV-1. N. Engl. J. Med. https://doi.org/10.1056/NEJMc2004973.

[To 2020] To, K.K.-W., Tsang, O.T.-Y., Leung, W.-S., Tam, A.R., Wu, T.-C., Lung, D.C., Yip, C.C.-Y., Cai, J.-P., Chan, J.M.-C., Chik, T.S.-H., Lau, D.P.-L., Choi, C.Y.-C., Chen, L.-L., Chan,W.-M., Chan, K.-H., Ip, J.D., Ng, A.C.-K., Poon, R.W.-S., Luo, C.-T., Cheng, V.C.-C.,Chan,J.F.-W.,Hung,I.F.-N.,Chen,Z.,Chen,H.,Yuen,K.-Y.,2020.Temporal profiles of viral load in posterior oropharyngeal saliva samples and serum antibody responses during infection by SARS-CoV-2: an observational cohort study. Lancet. Infect. Dis. https://doi.org/10.1016/S1473-3099(20)30196-1

[Pan 2020] Pan, Y., Zhang, D., Yang, P., Poon, L. L. M., & Wang, Q. (2020). Viral load of SARS-CoV-2 in clinical samples. The Lancet Infectious Diseases, 20(4). https://doi.org/10.1016/S1473-3099(20)30113-4

[Watanabe 2010] T. Watanabe, T.A. Bartrand, M.H. Weir, T. Omura, C.N. Haas. Development of a dose-response model for SARS coronavirus Risk Anal., 30 (2010), pp. 1129–1138, 10.1111/j.1539-6924.2010.01427.x

[Mueller 2020] Amy V Mueller, Matthew J. Eden, Jessica J. Oakes, Chiara Bellini, Loretta A Fernandez Quantitative Method for Comparative Assessment of Particle Filtration Efficiency of Fabric Masks as Alternatives to Standard Surgical Masks for PPE edRxiv 2020.04.17.20069567; doi: https://doi.org/10.1101/2020.04.17.20069567

[Li 2007] Li Y, Leung GM, Tang JW, Yang X, Chao CY, et al. (2007) Role of ventilation in airborne transmission of infective agents in the built environment—A multidisciplinary systematic review. Indoor Air 17: 2–18.

[Marr 2012] Marr, David; Mason, Mark; Mosley, Ron; Liu, Xiaoyu. 2012 The influence of opening windows and doors on the natural ventilation rate of a residential building. The Free Library (January, 1), https://www.thefreelibrary.com/The influence of opening windows and doors on the natural ventilation…- a0282940533

[Abuhegazy 2020] M. Abuhegazy1 e et al. (2020) Num. investigation of aerosol transport in a classroom with relevance to COVID-19 Physics of Fluids 32, 103311, https://doi.org/10.1063/5.0029118

